# Image transmission through a multimode fibre in reflection mode with physics-guided deep learning towards ultrathin endoscopy

**DOI:** 10.64898/2026.08.28.26361674

**Authors:** Ziyu Ye, Feng He, Tianrui Zhao, Wenfeng Xia

**Author notes:** Author to whom any correspondence should be addressed. **E-mail:**.

## Abstract

Ultrathin endoscopy is highly attractive for real-time tissue imaging in narrow and hard-to-reach regions of the body. A single multimode fibre (MMF) is an attractive probe because of its small diameter, flexiblility, and diffraction-limited spatial resolution enabled by the large number of transverse modes guided within a single core. Because the distal fibre tip is inaccessible during endoscopy, reflection-mode imaging, in which the same fibre delivers illumination and collects backscattered light, is more practical than transmission-mode imaging. However, image recovery from the resulting speckle pattern is challenging because light undergoes double-pass propagation through the MMF, with mode coupling and dispersion; the backscattered signal is weak, and the camera records intensity only, without phase information. Here, we propose a single-shot reflection-mode MMF imaging framework that combines a reflected real-valued intensity transmission matrix (reflected-RVITM) with an image restoration network. The reflected-RVITM is calibrated using intensity-only measurements, without interferometry or phase retrieval, and provides a physics-guided initial reconstruction from a single backscattered speckle frame. A restoration network then refines this initial reconstruction instead of inverting the raw speckle. Four restoration backbones are evaluated: HPM-Attention-UNet, GAM, MambaIRv2, and CICPNet. On matched datasets, hybrid models outperformed corresponding networks trained to map raw speckle directly to images. For example, HPM-Attention-UNet on MNIST improved mean PCC from 0.572 to 0.944 (+65.1%). Under domain shift, with training only on Fashion-MNIST and tested on unseen CIFAR scenes, hybrid models achieved mean PCC of 0.61–0.65, compared with 0.36–0.50 for direct learning. This framework is further demonstrated using physical objects at the distal fibre tip. These results demonstrate that a reflected-RVITM physics prior combined with a restoration network enables single-shot image recovery after intensity-only calibration, offering a phase-retrieval-free and generalisable route towards minimally invasive reflection-mode MMF endoscopy.

## 1 Introduction

Endoscopy is a key tool for minimally invasive diagnosis and image-guided intervention [1, 2]. It enables clinicians to inspect internal tissues through small incisions or natural orifices with minimal disruption to surrounding anatomy. However, many clinically important targets are located in narrow and hard-to-reach sites, such as the pancreaticobiliary ducts, small airways, deep brain regions, and needle-accessible lesions. Reaching these sites safely requires probes that are both ultrathin and highly flexible. Conventional miniature endoscopes typically rely on distal optics, relay elements, scanning mechanisms, each of which imposes trade-offs among probe diameter, flexibility, and image performance and system complexity [3, 4]. Thin optical fibres therefore offer a compelling alternative for realising ultrathin, flexible endoscopes with the potential to overcome these limitations [5–8].

Coherent fibre bundles (CFB) and multimode fibres (MMFs) are two of the widely used probe architectures for ultrathin endoscopy. In a coherent fibre bundle, the image is sampled by discrete core array samples, leading to characteristic honeycomb pixelation artefacts and a spatial resolution fundamentally limited by the core pitch [9]. In contrast, MMF-based endomicroscopy offers several significant advantages over CFB-based systems, notably: disposable fibre costs reduced by 2–3 orders of magnitude, and 1–2 orders of magnitude higher spatial resolution [7, 10]. These advantages, however, come at the cost of increased computational complexity. Mode dispersion and coupling scramble the input into a seemingly random speckle pattern at the fibre output [11], making direct image formation impossible. Consequently, the object image must be computationally reconstructed from the measured speckle intensity pattern.

Existing reconstruction approaches can be broadly divided into two categories. The first relies on fiber calibration, typically through transmission matrix measurements [12, 13], digital phase conjugation [14], or compressed sensing [15]. Many of these field-based approaches require interferometric phase measurement, and their performance is highly sensitive to fibre bending and environmental perturbations that alter the calibrated transmission characteristics. A second category uses deep learning to directly map the measured speckle to object image [16–19]. Although these methods eliminate the need for phase measurement, they typically require large, representative training datasets and often exhibit limited generalisation to imaging conditions outside of the training distribution. Furthermore, the vast majority of learning-based methods have been demonstrated only in transmission-mode imaging, where the object is located beyond the distal facet of the fibre [12, 13, 16, 17].

Reflection-mode imaging is more compatible with practical endoscopy, where access to the distal end of the fibre is unavailable during clinical use. In this configuration, a single MMF both delivers illumination to the tissue and collects the backscattered light, enabling imaging through proximal-end access alone. However, image reconstruction is considerably more challenging because the optical field propagates through the fibre twice, the backscattered signal is inherently weak, and only the speckle intensity is measured at the proximal end. Recovering the object image from the reflected speckle pattern remains a significant challenge. Liu *et al*. demonstrated single-shot wide-field reflectance imaging through a single MMF using a learning-assisted reflection matrix approach, and achieved imaging of real objects [20]. More recently, deep networks have improved the robustness of single-MMF imaging to fibre bending [21] and temperature variations [22]. However, these approaches either rely on learning-assisted calibration or inversion of the reflected speckle response, or are designed to address specific perturbations. Whether an intensity-only, calibration-based physics prior, combined with a separate image-domain restoration step, can provide robust and generalisable reconstruction for reflection-mode MMF imaging remains open.

To address these challenges, we build on the real-valued intensity transmission matrix (RVITM), an intensity-only calibration framework previously developed by our group for transmission-mode MMF imaging [23, 24]. Unlike conventional transmission-matrix approaches, the RVITM establishes an approximately linear mapping between the input and output intensities, eliminating the need for interferometric phase measurements [23]. Here, we extend this framework to reflection mode MMF imaging by calibrating the reflected speckle response directly under the same double-pass geometry, thereby reconstructing a reflected RVITM that empirically models the intensity-to-intensity relationship of the reflection channel. The reflected RVITM provides an initial reconstruction from a single speckle frame, after which a restoration network refines this initial reconstruction rather than directly inverting the raw speckle. This decouples the physics-based reconstruction from the data-driven restoration stage, allowing the network to operate in the image domain instead of the highly complex speckle domain. We therefore investigate whether restoring initial reconstructions, rather than directly reconstructing from the raw speckle patterns, improves generalisation to previously unseen image types and real objects. To further examine the importance of long-range modelling in this restoration stage, we combine the reflected RVITM with several network backbones, including attention-, state-space-, and convolutional-attention-based architectures [21, 24–26].

## 2 Methods

### 2.1 Reflection-mode MMF imaging system

The reflection-mode single-MMF imaging system is shown in Figure 1(a). Illumination was provided by a continuous-wave laser (532 nm, 50 mW; PGL-FC-532nm-50mW, CNI Laser, Changchun, China). A mounted zero-order half-wave plate (HWP; WPH10M-532, Thorlabs, Newton, NJ, USA) and a cage-cube polarising beam splitter (PBS; CCM1-PBS251/M, Thorlabs) controlled the laser power and polarisation. The beam was expanded and collimated by a telescope formed by a mounted achromatic doublet L1 (50 mm effective focal length; AC254-50-A-ML, Thorlabs) and a mounted achromatic doublet L2 (200 mm; ACT508-200-A-ML, Thorlabs), redirected by a mirror (M), and focused by a mounted achromatic doublet L3 (75 mm; ACT508-75-A-ML, Thorlabs) onto a cage-cube non-polarising beam splitter (BS; CCM1-BS013/M, Thorlabs). The BS coupled the beam into the proximal facet of a step-index multimode fibre (MMF; FG600LEA, Thorlabs; 600 µm core diameter, 0.22 numerical aperture, low-OH, 2 m length).

**Figure 1.**
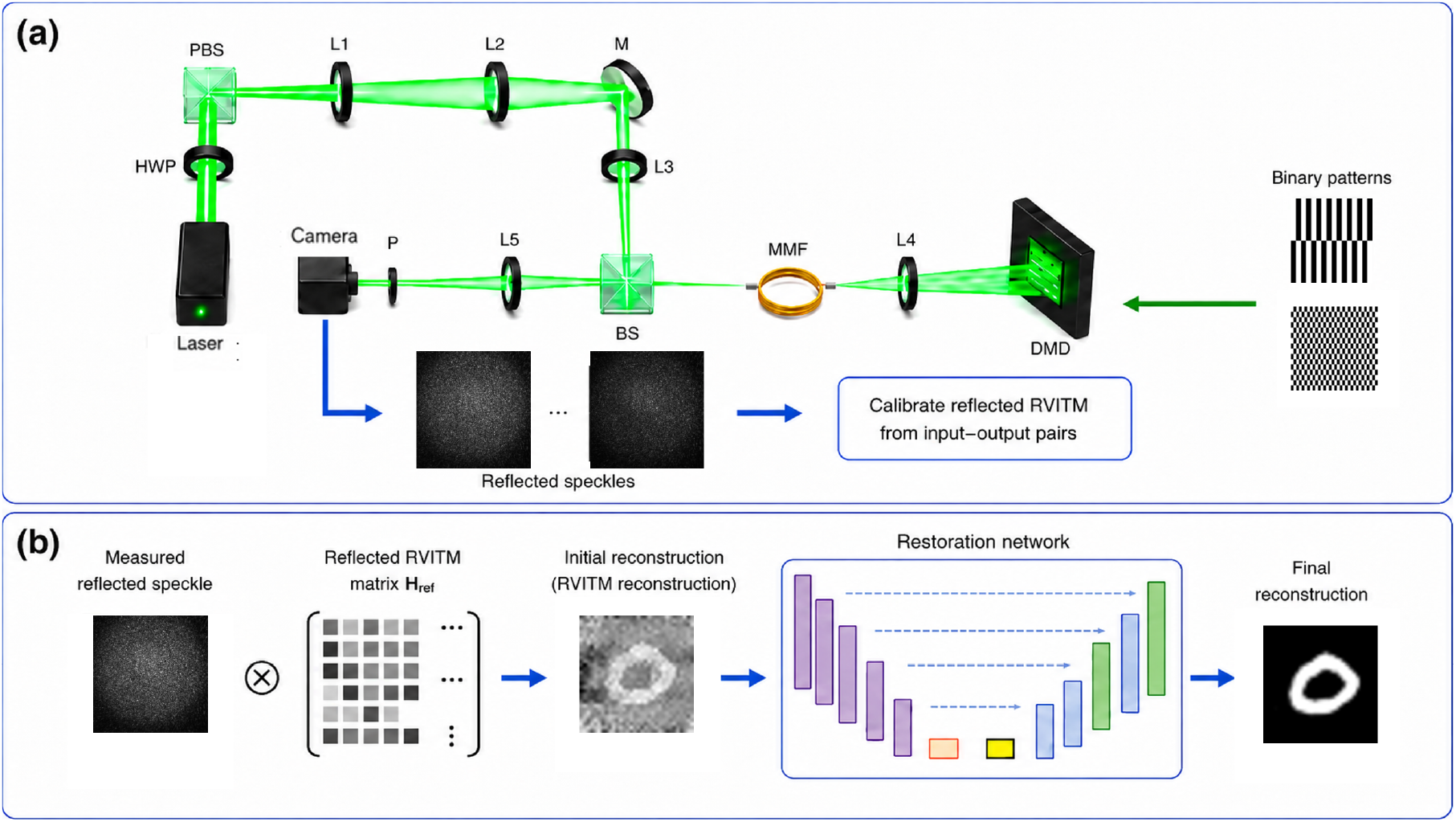
Reflection-mode single-MMF imaging framework. (a) Optical system: the laser beam (532 nm) is conditioned by a half-wave plate (HWP) and a polarising beam splitter (PBS), expanded and collimated by achromatic doublets L1 (50 mm) and L2 (200 mm), redirected by mirror M, and focused by L3 (75 mm) through a non-polarising beam splitter (BS) onto the proximal facet of the multimode fibre (MMF). At the distal end, L4 (50 mm) relays the fibre output onto the target plane. For controlled experiments, a digital micromirror device (DMD) displays calibration patterns or target images. Reflected light is collected by the same MMF and returned to the proximal end, separated from the illumination path by the BS, and relayed by L5 (50 mm) through a polariser (P) onto the camera to record the reflected speckle intensity. (b) Reconstruction workflow: the measured reflected speckle pattern is first mapped by the reflected-RVITM matrix **R**_ref_ to generate a physics-guided initial reconstruction, which is subsequently refined by a restoration network to obtain the final image.

At the distal facet, a mounted achromatic doublet L4 (50 mm; AC254-50-A-ML, Thorlabs) projected the fibre output onto a digital micromirror device (DMD; DLP7000, Texas Instruments, Dallas, TX, USA), which displayed binary patterns. The light reflected from the DMD was collected by the same MMF and propagated back to the proximal end. The returned light was separated from the illumination path by the BS and relayed by a mounted achromatic doublet L5 (50 mm; AC254-50-A-ML, Thorlabs) through a polariser (P; P-LPVISE100-A, Thorlabs) onto a CMOS camera (C11440-22CU01, Hamamatsu Photonics, Hamamatsu, Japan), which recorded the reflected speckle intensity.

Figure 1(b) illustrates the reconstruction pipeline following calibration. For each target, a single reflected speckle pattern is recorded by the camera. The reflected RVITM (see details in section 2.2) first maps the measured reflected speckle pattern to a physics-guided initial reconstruction. A restoration network (section 2.3) then suppresses reconstruction artifacts and refines this initial estimate to produce the final reconstruction. During imaging, the entire reconstruction is performed from a single camera frame, eliminating the need for raster scanning. In this study, the distal target was either a DMD-generated binary pattern for controlled evaluation or a physical object placed at the distal end for practical imaging tests, while all measurements were acquired exclusively from the proximal end.

### 2.2 Reflected real-valued intensity transmission matrix (reflected-RVITM)

In reflection mode, light undergoes a double pass through the same MMF, propagating from the proximal end to the distal target plane and returning to the proximal camera after interacting with the target. Mode coupling and dispersion scramble the optical field during each pass, and the forward and backward propagation can be described by complex-valued transmission matrices (TMs), **U**_f_ and **U**_b_, respectively, which map optical fields to optical fields while preserving both amplitude and phase information [12]. To reduce system complexity, we adopt the real-valued intensity transmission matrix (RVITM) framework [23], in which the camera records only the intensity of the reflected speckle pattern. The double-pass optical channel is therefore represented by the reflected-RVITM **R**_ref_, which provides an intensity-to-intensity mapping between the distal target and the measured proximal speckle pattern.

#### Double-pass field model

The proximal illumination is fixed during acquisition. Let **e**_0_ ∈ℂ^*N*^denote the corresponding proximal input field, where *N* = *N*_*x*_*N*_*y*_, with *N*_*x*_ and *N*_*y*_ denoting the numbers of spatial sampling points along the *x*- and *y*-directions, respectively. Let **U**_f_ ∈ℂ^*N* ×*N*^denote the forward TM. The resulting distal field is

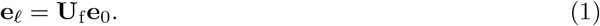

Let ***ρ*** ∈ℂ^*N*^denote the complex reflectance on the distal plane. Target-independent distal efficiency factors, including the fixed reflection and coupling efficiency, DMD diffraction efficiency, and collection efficiency, are incorporated into a fixed complex diagonal gain matrix **D**_g_ ∈ ℂ^*N* ×*N*^. The field returning into the MMF is then given by

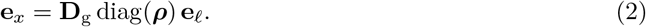

Let **U**_b_ ∈ℂ^*M* ×*N*^denote the backward TM, where *M* is the number of camera sampling points. The returning field at the proximal end is:

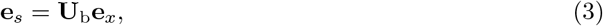

and the camera records the corresponding speckle intensity vector ***s*** = |**e**_*s*_| ^*⊙*2^∈ℝ^*M*^. Combining equations (1)–(3) gives the coherent double-pass map

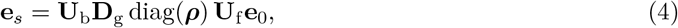

#### Reflected-RVITM approximation

The RVITM approximation replaces the coherent field model in (1)–(4) with an intensity-only representation. Specifically, squared magnitudes are taken at each stage, while coherent interference terms associated with **U**_f_, **U**_b_, and diag(***ρ***) are neglected. Each propagation step is therefore represented by an intensity-domain transmission matrix.

Because the proximal illumination is fixed during acquisition, we use a uniform reference intensity vector **i**_0_ = vec(**1**) ∈ℝ^*N*^, with spatial non-uniformity of the actual illumination absorbed into the forward intensity transmission matrix. The distal illumination intensity is then approximated as

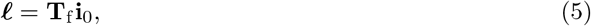

where ***l*** =|**e**_*l*_| ^*⊙*2^denotes the distal illumination intensity |**U**_f_**e**_0_| ^*⊙*2^and **T**_f_ ∈ℝ^*N* ×*N*^is the forward intensity transmission matrix. Let ***p*** = |***ρ***| ^*⊙*2^ denote the intensity reflectance at the distal plane, and let ***g*** contain the squared magnitudes of the diagonal entries of **D**_g_. Neglecting the phase of **D**_g_, the intensity of the returning field at the distal plane is then approximated by:

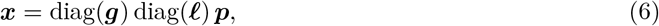

corresponding to equation (2). The proximal speckle intensity is then approximated by

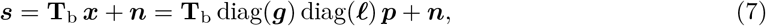

where **T**_b_ ∈ℝ^*M* ×*N*^ is the backward intensity transmission matrix and ***n*** is measurement noise. For a fixed optical alignment, ***l*** and ***g*** remain constant. Their combined effect with the backward intensity transmission matrix can therefore be absorbed into a single reflected RVITM,

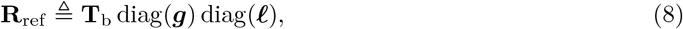

giving

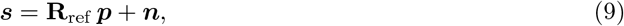

where **R**_ref_ ∈ℝ^*M* ×*N*^ is the reflected RVITM and *M* denotes the number of speckle pixels. Equations (5), (6), (7), (8) and (9) summarise the intensity-only approximation used thereafter.

#### Calibration

Following the RVITM calibration procedure [23], the reflected RVITM **R**_ref_ is calibrated using a Hadamard matrix **H** ∈ *{*−1, +1 *}*^*N* ×*N*^(*N* = 32 × 32 = 1024). Since the DMD supports only binary 0*/*1 modulation, **H** is decomposed into complementary binary patterns

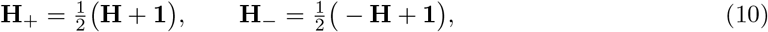

where **1** ∈ℝ^*N* ×*N*^is an all-ones matrix (equation (10)). During calibration, the distal intensity reflectance ***p*** is sequentially set to the binary DMD patterns given by the columns of **H**_+_ and **H** . One reflected speckle frame is recorded for each column, yielding the measurement matrix **S** ∈ℝ^*M* ×2*N*^. Let ***s***_1_ denote the response to the all-ones pattern, corresponding to the first column of **H**_+_. Subtracting this reference response from each measured frame and converting the measurements back to the *±*1 Hadamard basis, the reflected RVITM is recovered using the orthogonality relation [**H**, −**H**][**H**, −**H**]^⊤^= 2*N* **I**:

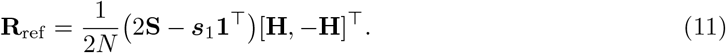

#### Reconstruction

For an unknown target, the reflected speckle intensity ***s*** recorded by the camera is inverted using the pseudoinverse of the calibrated reflected RVITM,

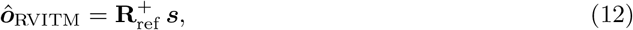

where 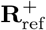 denotes the Moore–Penrose pseudoinverse of **R**_ref_ (equation (12)). The resulting vector *ô*_RVITM_ provides an estimate of the target intensity reflectance ***p***. It is subsequently reshaped into a two-dimensional image to form the initial reconstruction, which is then passed to the restoration network described in Section 2.3.

### 2.3 Restoration networks

The restoration networks take the initial reconstruction *ô*_RVITM_ as input and generate a refined image *ô*. This formulation converts the original reflected-speckle inversion problem into an image-domain restoration task. Since the reflected-RVITM stage provides a physics-guided initial reconstruction, the network is mainly required to suppress residual speckle-like artifacts, enhance contrast, and recover fine structural details.

Four restoration backbones were evaluated, as summarised in Table 1. CICPNet is a convolutional attention U-Net designed for reflected MMF speckle reconstruction [21].

**Table 1.** Restoration backbone configurations and parameter counts.

| Backbone | Core mechanism | Configuration | Params |
| --- | --- | --- | --- |
| CICPNet [21] | Conv. attention U-Net | 5 levels, 64–1024 ch. | 28.63 M |
| HPM-Attention-UNet [24] | Multi-scale U-Net + skip attention | 4 levels, 64–512 ch. | 5.53 M |
| GAM [25] | Residual U-Net + global attention | 4 levels, 16–256 ch. | 5.18 M |
| MambaIRv2 [26] | State-space model + local attention | 4 stages, embed 96 | 4.93 M |

HPM-Attention-UNet is a hierarchical parallel multi-scale attention U-Net previously developed for RVITM-assisted MMF image retrieval [24]. GAM is a residual U-Net incorporating global attention modules designed to capture non-local channel–spatial dependencies in image transmission through MMFs and turbid media [25]. MambaIRv2 is a state-space image restoration model that combines selective state-space modelling with local attention for efficient long-range feature extraction [26].

### 2.4 Datasets and experimental protocol

Three image datasets were used for training, validation and testing (Table 2): MNIST handwritten digits [27], Fashion-MNIST apparel images [28], and CIFAR natural scenes [29]. Each image was resized to 32 × 32 pixels and converted to a DMD-compatible target pattern. Each target pixel was mapped to a 4 × 4 block of DMD micromirrors centred on the active DMD region. For each target, a single reflected speckle pattern was recorded at the proximal camera, forming paired reflected-speckle and ground-truth images.

**Table 2.** Datasets used for training, validation, and testing. For each dataset, paired reflected-speckle and ground-truth images were acquired and split into the counts shown.

| Dataset | Train | Val | Test |
| --- | --- | --- | --- |
| MNIST | 7000 | 1000 | 2000 |
| Fashion-MNIST | 7000 | 1000 | 2000 |
| CIFAR | 7000 | 1000 | 2000 |

The reflected RVITM was calibrated once per fixed optical alignment and fibre state using the calibration procedure described in section 2.2. The calibrated matrix **R**_ref_ was then reused for all targets acquired under the same calibration. If the fibre state or optical alignment changed, the reflected RVITM was recalibrated using equation (11).

For each restoration backbone, two reconstruction strategies were evaluated. In the hybrid strategy, the network received the 32 × 32 initial reconstruction and predicted the refined 32 × 32 image. In the direct-learning baseline, the same backbone family was trained to map the raw reflected speckle directly to the same 32 × 32 ground truth image, without the reflected-RVITM stage. The hybrid and direct-learning variants used the same data splits, loss function, model-selection criterion, and optimisation protocol. Because the hybrid input is already a 32 × 32 image whereas the direct-learning input is a full-resolution reflected speckle, each direct-learning variant used an additional convolutional input stem that downsamples the speckle to the spatial resolution expected by the shared encoder; the remaining encoder–decoder stages and channel widths followed the same family configuration as the corresponding hybrid model (Table 1), so that the comparison isolates the effect of the reflected-RVITM front end rather than a change of backbone capacity.

Two evaluation settings were used. In the matched-dataset setting, each model was trained, validated, and tested on images from the same dataset, allowing reconstruction performance to be assessed under in-distribution conditions. In the cross-dataset setting, models were trained on Fashion-MNIST and tested on the held-out CIFAR test set. Model selection and early stopping used only the Fashion-MNIST validation split; no CIFAR image was used during training or checkpoint selection. This setting was designed to assess generalisation under a deliberate domain shift from apparel images to unseen natural scenes.

Finally, to assess the practical imaging capability of the proposed framework beyond dataset-based evaluation, the calibrated system was tested on previously unseen physical objects placed at the distal end of the MMF. These objects were completely independent of the image datasets and were not used for training, validation, or model selection. The corresponding reflected speckle patterns were acquired experimentally and processed using the same calibrated reflected RVITM and trained restoration network, without any target-specific recalibration or fine-tuning.

This experiment provides an independent validation of the end-to-end reconstruction pipeline and assesses its ability to transfer from digitally generated target patterns to real physical objects, thereby demonstrating the practical feasibility of MMF-based reflection-mode imaging.

All models were trained using the mean-squared-error (MSE) loss between the network output and the ground-truth image for up to 200 epochs with a cosine learning-rate schedule. Checkpoint selection used the lowest validation MSE; validation PCC and SSIM were monitored as secondary diagnostics to confirm that the selected checkpoint did not trade structural fidelity for a lower MSE alone. Within each backbone family, the hybrid and direct-learning variants shared the same optimiser (Adam or AdamW, chosen once per family), the same initial learning rate (selected once per family from the range 1× 10^−4^–1 × 10^−3^on the validation split), and the same maximum number of training epochs. The batch size was set to the largest value that fitted on a single NVIDIA A100 GPU for the corresponding input size and was matched within each hybrid/direct pair whenever memory permitted.

### 2.5 Evaluation metrics

Reconstruction quality was quantified using the Pearson correlation coefficient (PCC) and structural similarity index measure (SSIM), which provide complementary measures of image fidelity. PCC measures the linear correlation between the reconstructed and ground truth images and is insensitive to global intensity scaling and offset. SSIM evaluates structural similarity by jointly comparing local luminance, contrast, and structural information.

For a reconstruction *ô* and ground truth ***o***, PCC was defined as

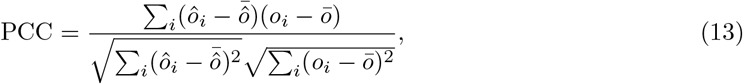

where 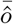 and *ō*denote the mean intensities of the reconstruction and ground truth, respectively.

SSIM was defined as

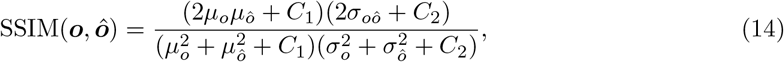

where *µ*_*o*_ and *µ*_*ô*_are local mean intensities, 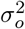 and 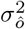 are local variances, *σ*_*oô*_ is the local covariance, and *C*_1_ and *C*_2_ are stabilising constants.

Both metrics (13) and (14) were computed for each test image against the corresponding ground truth. Unless stated otherwise, reported values in the text are test-set means. The full per-image score distributions are shown as box plots (Figures 2, 4 and 7; box-plot conventions are defined in those figure captions). The hybrid and direct-learning variants of each backbone were evaluated on the same test images, yielding paired per-image scores. Pairwise improvements of hybrid over direct learning were tested with two-sided paired Student *t*-tests on the per-image differences. Reported *p*-values are Holm–Bonferroni-adjusted across the four backbones within each protocol (matched or cross-dataset) × dataset × metric family. Relative changes are quoted as percentages of the corresponding direct-learning mean.

**Figure 2.**
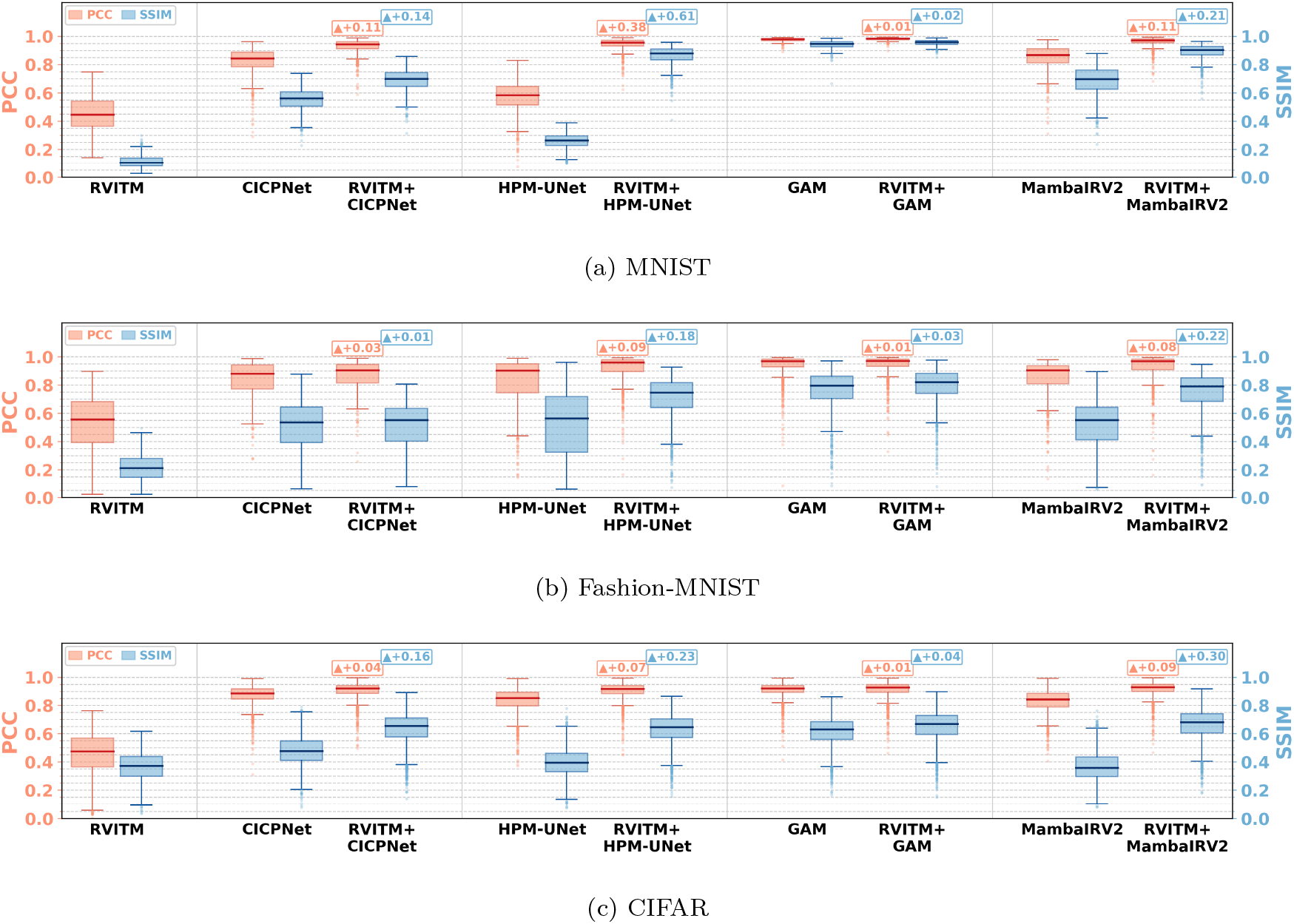
Matched-dataset quantitative comparison of per-image PCC and SSIM. Within each method cluster, the left box (and left vertical axis) is PCC and the right box (and right vertical axis) is SSIM. In each box, the central line is the median, the box spans the first to third quartiles (Q1–Q3; interquartile range IQR = Q3 −Q1), and the whiskers extend to the most extreme observations still within 1.5 × IQR of the box (Tukey fence); individual points beyond the whiskers denote outliers. Sample minima and maxima therefore appear either as whisker ends (if within the fence) or as outliers (if outside it). Columns compare the RVITM baseline, direct learning and hybrid RVITM-guided restoration for each backbone (direct models use the backbone name; hybrids are labelled RVITM+backbone). Annotated values are paired mean differences relative to direct learning.

## 3 Results

### 3.1 Matched-dataset reconstruction

In the matched-dataset evaluation (section 2.4), each model was trained, validated, and tested on the same dataset: MNIST, Fashion-MNIST, or CIFAR. Three reconstruction strategies were compared: the analytic initial reconstruction alone, direct speckle-to-image learning, and the proposed hybrid strategy in which a restoration network refines the initial reconstruction. The direct-learning and hybrid variants used the same four backbone families: CICPNet, HPM-Attention-UNet, GAM, and MambaIRv2. Figure 2 shows per-image PCC and SSIM distributions as box plots, and Figure 3 shows representative reconstructions. Across all three datasets, the initial reconstruction recovered the coarse spatial organisation of the target but exhibited substantial speckle-like artifacts, reduced contrast, and limited structural fidelity. Relative to this training-free RVITM baseline (mean PCC 0.45–0.51 and mean SSIM 0.11–0.36 across the three datasets), every matched pure-DL and hybrid model increased both metrics (Figure 2). Across backbone–dataset combinations, the hybrid RVITM-guided models further outperformed their direct-learning counterparts: for every backbone and metric except one case noted below, the paired hybrid-minus-direct mean difference was positive with Holm–Bonferroni-adjusted *p <* 0.001.

**Figure 3.**
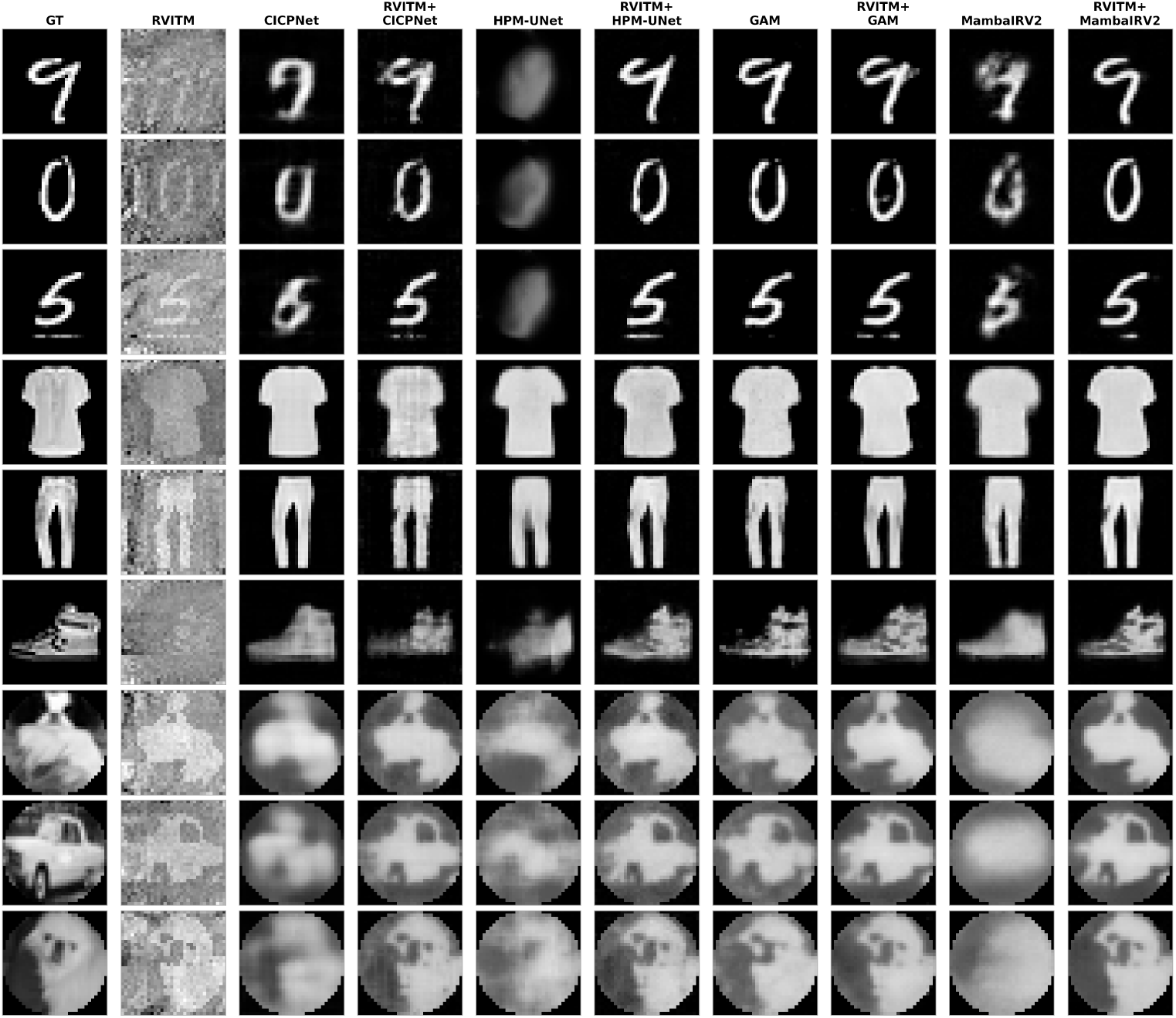
Matched-dataset visual examples. Rows show example reconstructions from MNIST, Fashion-MNIST, and CIFAR. Columns show the ground truth, the initial (RVITM) reconstruction, and each backbone (CICPNet, HPM-Attention-UNet, GAM, and MambaIRv2) as direct-learning and RVITM+backbone hybrid variants.

The magnitude of the RVITM guidance benefit depended on the strength of the direct-learning baseline, which is visible both in the means and in the box-plot spreads. The largest gains occurred when direct learning was weak. For HPM-Attention-UNet on MNIST, mean PCC rose from 0.572 (direct) to 0.944 (hybrid; +65.1%) and mean SSIM from 0.260 to 0.864 (+231.7%; both paired *t*-tests, Holm–Bonferroni-adjusted *p <* 0.001). The corresponding PCC boxes shifted from a median of 0.584 (Q1–Q3: 0.514–0.646; min–max: 0.074–0.829) under direct learning to a median of 0.956 (Q1–Q3: 0.932–0.971; min–max: 0.623–0.990) for the hybrid model, so that the hybrid interquartile range lay entirely above the direct-learning upper quartile. Comparable large SSIM gains were obtained for MambaIRv2 on MNIST (mean from 0.686 to 0.889; +29.7%) and on CIFAR (from 0.371 to 0.661; +78.3%). When direct learning already achieved near-saturated PCC, absolute hybrid gains were small. For GAM on CIFAR, mean PCC increased only from 0.909 to 0.912 (+0.3%), while SSIM improved from 0.614 to 0.650 (+5.9%); both paired hybrid-versus-direct differences remained positive after Holm adjustment (*p <* 0.001). Hybrid–hybrid differences on matched CIFAR PCC remained small (means 0.904–0.915) relative to the hybrid-versus-direct margin. The sole non-significant hybrid-versus-direct comparison was Fashion-MNIST SSIM for CICPNet (from 0.510 to 0.515; Holm–Bonferroni-adjusted *p* = 0.25), where the absolute gain was within 1%.

Overall, these results show that the reflected-RVITM stage provides a useful physics-guided prior for subsequent image restoration. Its contribution is most pronounced when direct speckle-to-image learning struggles to recover the underlying target structure, while remaining beneficial—and statistically significant for PCC—for strong restoration backbones that already achieve high in-distribution performance.

### 3.2 Cross-dataset generalisation from Fashion-MNIST to CIFAR

To evaluate generalisation under domain shift, all learned models were trained exclusively on Fashion-MNIST and tested on the held-out CIFAR test set. The training-free RVITM initial reconstruction was included as a reference.

Figure 4 summarises cross-dataset PCC and SSIM, and Figure 5 shows representative reconstructions on unseen CIFAR scenes.

**Figure 4.**
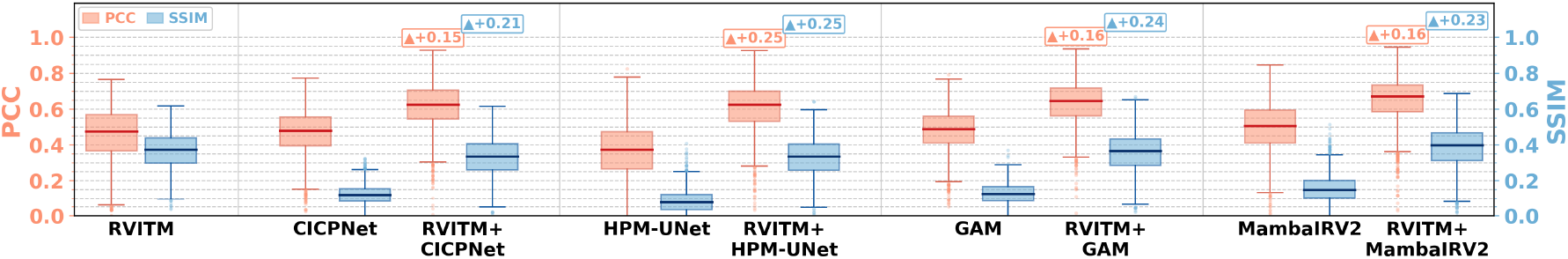
Cross-dataset quantitative comparison. Models were trained on Fashion-MNIST and tested on unseen CIFAR images. Within each method cluster, the left box (and left vertical axis) is PCC and the right box (and right vertical axis) is SSIM. In each box, the central line is the median, the box spans Q1–Q3 (IQR = Q3 −Q1), and Tukey whiskers extend to the extreme values within 1.5 × IQR; points beyond the whiskers are outliers. Pairwise mean changes relative to the direct-learning baseline are annotated on the plot.

**Figure 5.**
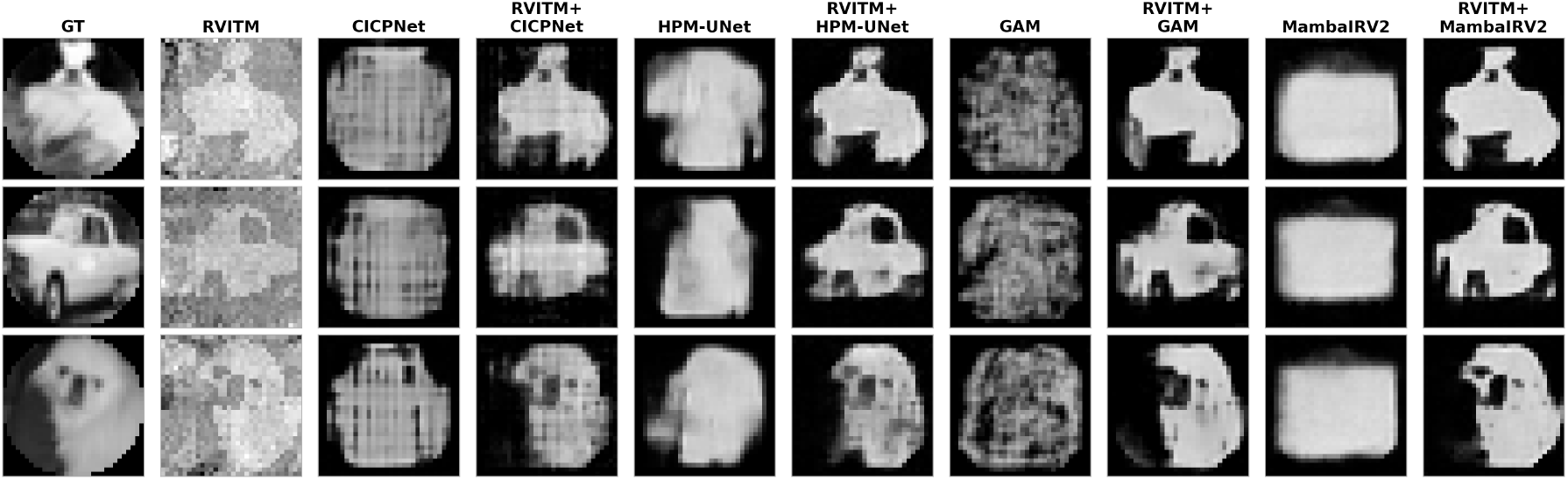
Cross-dataset visual examples. Models were trained on Fashion-MNIST and tested on unseen CIFAR images. Rows show three example CIFAR test images. Columns show the ground truth, the initial (RVITM) reconstruction, and each backbone (CICPNet, HPM-Attention-UNet, GAM, and MambaIRv2) as direct-learning and RVITM+backbone hybrid variants.

Compared with matched CIFAR testing, direct-learning models exhibited a large domain-shift penalty under Fashion-MNIST *→*CIFAR transfer. Mean PCC on CIFAR fell from 0.83–0.91 (matched pure-DL baselines) to 0.36–0.50 (cross pure-DL), corresponding to absolute drops of about 0.33–0.47. Mean SSIM fell from 0.37–0.61 with matched to 0.08–0.15 under transfer, falling below the training-free RVITM initial reconstruction for all four backbones. This degradation indicates that direct speckle-to-image learning was strongly dependent on the training image distribution and struggled to generalise to the unseen natural-scene domain.

In contrast, the hybrid models were substantially more robust to the same domain shift. All four hybrid backbones outperformed their direct-learning counterparts, with mean PCC in the range 0.61–0.65 for hybrids versus 0.36–0.50 for direct learning (Figure 4; paired *t*-tests of hybrid versus direct learning, Holm–Bonferroni-adjusted *p <* 0.001 for both PCC and SSIM on every backbone). Relative PCC gains under transfer ranged from +30.5% (MambaIRv2) to +67.4% (HPM-Attention-UNet). RVITM+MambaIRv2 and RVITM+GAM achieved the highest cross-dataset mean PCC (0.649 and 0.635, respectively); differences among hybrids were much smaller than the hybrid-versus-direct margin. Compared with the training-free RVITM reconstruction, hybrid models improved PCC on the held-out CIFAR set, while residual SSIM variation across backbones remained visible. These results suggest that the principal advantage of the hybrid strategy under domain shift is the preservation and refinement of the coarse spatial structure already encoded by the physics-guided initial reconstruction.

The qualitative results in Figure 5 support this interpretation. Direct-learning models tend to impose Fashion-MNIST-like priors on unseen natural scenes, producing garment-like textures, blurred object blobs, or nearly featureless outputs. In contrast, the hybrid models retain the coarse layout of the CIFAR objects because the inputs already contain coarse structure from the initial reconstruction. This behaviour was particularly evident for RVITM+MambaIRv2 and RVITM+GAM.

### 3.3 Real-object reflection-mode imaging

The calibrated system was further tested using physical objects placed at the distal end of the MMF, including a metal ventilation grille (Supplementary Video 1), a needle (Supplementary Video 2), and a printed letter (Supplementary Video 3). These objects were not included in any training set and differ from the DMD-displayed targets because their reflected signals depend on physical surface reflectance, scattering, and coupling conditions. Since pixel-aligned ground truth was not available, the results were assessed qualitatively and then quantified against a photograph-derived silhouette pseudo-ground truth.

Qualitatively, the initial RVITM reconstructions recovered the approximate outlines of the physical objects despite pronounced speckle-like artifacts (Figure 6). The direct-learning models trained on Fashion-MNIST frequently suppress the object or introduce image features inconsistent with the physical targets. In contrast, the hybrid models retained the object-dependent structure present in the RVITM initial reconstruction while suppressing residual artifacts, enabling clearer recovery of the grille apertures, needle shaft, and letter strokes. Importantly, each reconstruction was obtained from a single camera frame without target-specific recalibration or fine-tuning. The corresponding frame-by-frame results are provided in Supplementary Videos 1–3.

**Figure 6.**
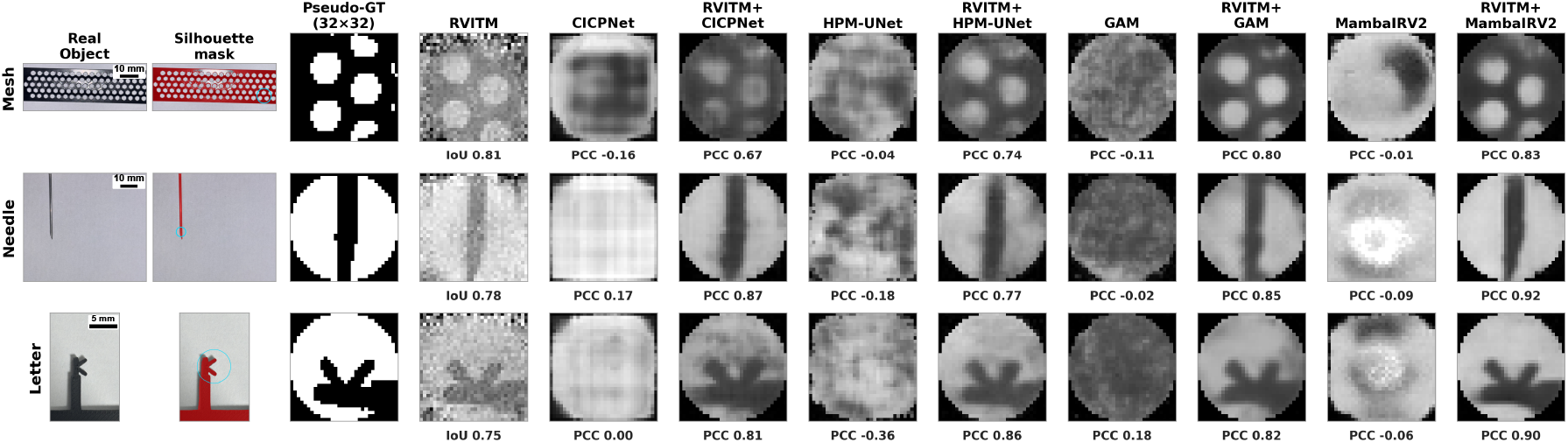
Real-object reflection-mode imaging. The first three columns are the real-object photograph, the segmented silhouette mask, and the 32 × 32 ground truth (GT). The remaining columns show one example frame per method. For RVITM, IoU against the GT measures alignment quality; the other methods report PCC. Scale bars: 10 mm (Mesh and Needle) and 5 mm (Letter). Supplementary Videos 1–3 show frame-by-frame reconstructions.

To provide a quantitative assessment, a silhouette pseudo-ground truth was constructed for each physical object from its photograph (Figure 6). A similarity transform aligning the photograph to the 32 × 32 reconstruction grid was fitted frame by frame, and only frames with RVITM–silhouette intersection-over-union (IoU) above 0.7 were retained so that subsequent Pearson correlation coefficient (PCC) scores primarily reflect reconstruction fidelity rather than registration error (Figure 7).

**Figure 7.**
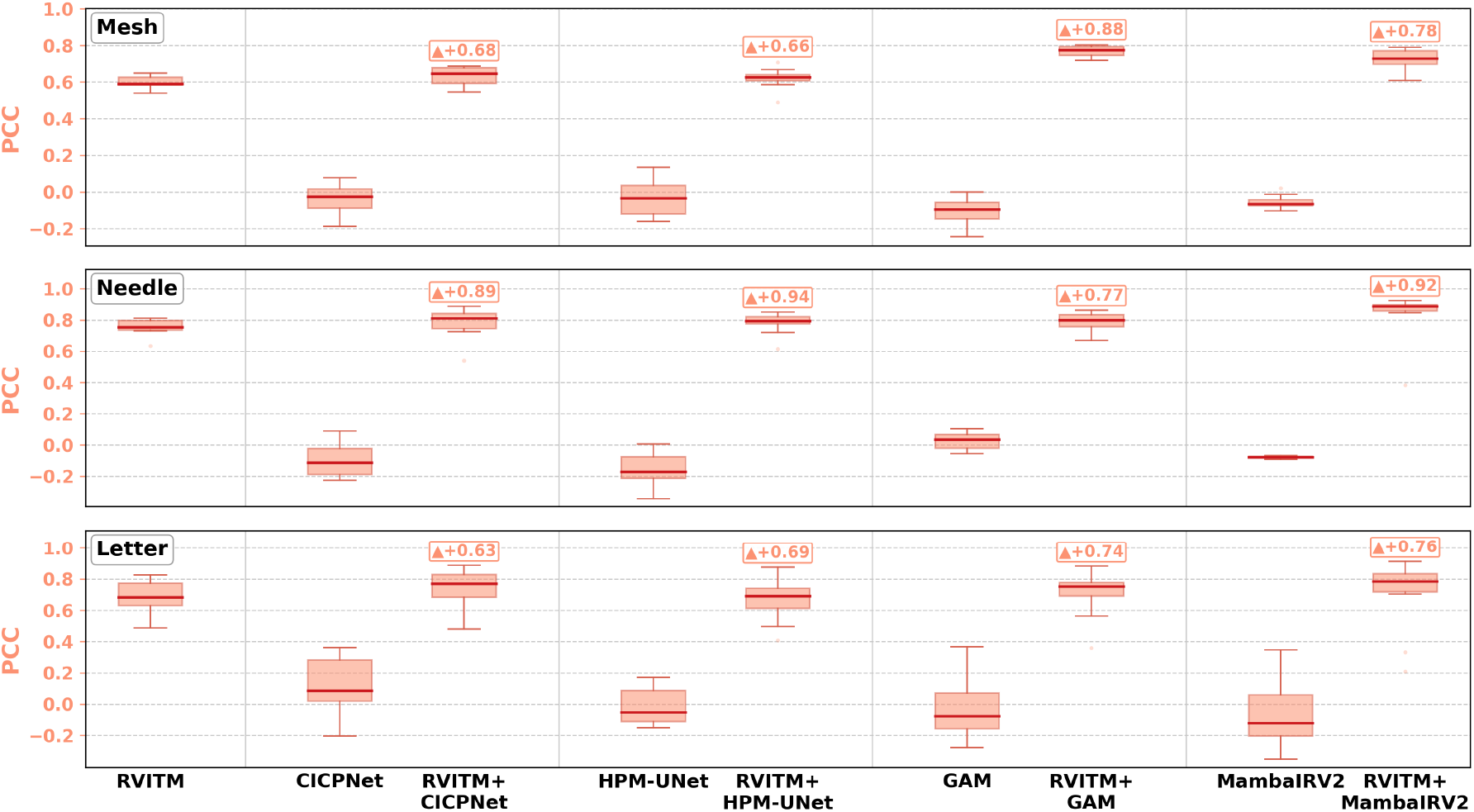
Real-object quantitative comparison. Per-frame PCC (single vertical axis) against the silhouette GT is shown for Mesh, Needle, and Letter, using frames in which the initial (RVITM) reconstruction achieved alignment IoU *>* 0.7. In each box, the central line is the median, the box spans Q1–Q3 (IQR = Q3 −Q1), and Tukey whiskers extend to the extreme values within 1.5 × IQR; points beyond the whiskers are outliers (PCC only, rather than dual-axis PCC/SSIM). Results are shown for the initial reconstruction baseline, direct-learning models, and hybrid RVITM-guided restoration models. For each backbone, the direct-learning model is labelled by the backbone name, whereas the hybrid model is labelled RVITM+backbone. Pairwise mean changes relative to the direct-learning baseline are annotated on the plot.

On the IoU-selected sequences (*n* = 92 Mesh frames, *n* = 182 Needle frames, *n* = 63 Letter frames), direct-learning mean PCC remained near zero or negative (for example Mesh direct-GAM −0.063; Needle direct-HPM-Attention-UNet −0.146), consistent with the qualitative failures of Fashion-MNIST-trained pure-DL models on physical targets. Hybrid models recovered substantially higher mean PCC: Mesh improved from 0.063 (GAM) to 0.775 (RVITM+GAM) and from −0.035 (MambaIRv2) to −0.700 (RVITM+MambaIRv2). Needle hybrids reached mean PCC 0.70–0.73 versus near-zero direct learning. Letter hybrids retained means near 0.67–0.69, close to the training-free RVITM baseline (0.687). These statistics are descriptive means over temporally correlated video frames and are not accompanied by formal *p*-values.

## 4 Discussion

We have demonstrated a single-shot reflection-mode single-MMF imaging framework in which an intensity-only reflected RVITM provides a physics-guided initial reconstruction from a single backscattered speckle frame, which is subsequently refined by an image-restoration network. On matched datasets, hybrid models improved mean PCC and SSIM over the corresponding direct-learning backbones with large, statistically significant gains when pure DL was weak (for example HPM-Attention-UNet on MNIST: PCC from 0.572 to 0.944; +65.1%). Under Fashion-MNIST *→* CIFAR transfer, hybrid mean PCC remained 0.61–0.65 while pure DL fell to 0.36–0.50 (paired *t*-tests, Holm–Bonferroni-adjusted *p <* 0.001). The same trained models also transferred to previously unseen physical objects placed at the distal fibre tip, where hybrid mean frame PCC on IoU-selected sequences substantially exceeded near-zero pure-DL scores.

The central advantage of the proposed framework arises from the separation of the optical inversion and image restoration tasks. The reflected-RVITM stage incorporates the measured double-pass optical response into an intensity-domain reconstruction, providing an object-dependent representation that is already spatially aligned with the target. The restoration network therefore does not need to learn the full mapping from a highly scrambled speckle pattern to an image; instead, it operates in the image domain to suppress residual artifacts and recover fine structure. This reduces the dependence of the learned mapping on the image statistics of the training set. In contrast, a direct speckle-to-image network must encode the entire optical inverse mapping within the network parameters, inherently coupling the reconstruction to the statistics of the training distribution. This distinction is particularly evident under domain shift: when trained on Fashion-MNIST and tested on unseen CIFAR scenes, the direct-learning models degraded sharply—notably in SSIM, where they fell well below the training-free initial reconstruction—whereas the hybrid models remained substantially more robust. The successful reconstruction of previously unseen physical objects further supports the ability of the physics-guided representation to transfer beyond the distribution of digitally generated training targets.

The choice of restoration backbone provided a secondary effect. MMF transmission distributes information from each object point across the output facet through modal interference, motivating architectures capable of modelling long-range spatial dependencies [24, 25]. Among the backbones evaluated, global attention (GAM) and selective state-space modelling (MambaIRv2) achieved the strongest cross-dataset performance. However, the reflected-RVITM front end remained the dominant factor: RVITM guidance improved all four backbone families, while the performance differences between backbones were comparatively smaller. These results suggest that the primary benefit comes from transforming the optical inverse problem into a physics-guided image-restoration problem, with architectural choices providing further but secondary gains.

Our approach shares the objective of single-shot reflection-mode single-MMF imaging with the learning-assisted reflection-matrix method of Liu *et al*. [20], but differs fundamentally in the role assigned to learning. Liu *et al*. learn an amplitude-to-amplitude mapping of the backscattered field directly from paired calibration and imaging data, whereas our approach first calibrates an intensity-only reflected RVITM and then applies learning exclusively in the image domain. The distinguishing contribution of the present work is therefore not a direct improvement in absolute in-distribution reconstruction quality, but the combination of phase-retrieval-free intensity calibration and a decoupled restoration stage that exhibits substantially improved robustness to domain shift and transfer to physical objects. This strategy is also complementary to recent learning-based approaches designed to compensate for specific perturbations, including mechanical bending [21] and temperature variations [22], which could potentially be incorporated into the proposed restoration framework.

Several limitations should be considered. The most important is calibration stability. The reflected RVITM is calibrated for a fixed fibre conformation and optical alignment, so substantial bending or temperature drift alters the double-pass response and degrades the calibrated matrix [11]. We did not characterise the window over which a single calibration remains valid. Moreover, although image acquisition itself is single-shot, recalibration requires sequential DMD patterns and is therefore not instantaneous. A promising direction is to combine the proposed framework with methods that improve tolerance to fibre perturbations. These include exploiting deformation-invariant features associated with the fibre memory effect [30], or training restoration networks across multiple fibre states to learn perturbation-tolerant representations [21, 22, 31].

Such approaches could reduce the calibration burden while retaining the modularity of the reflected-RVITM front end.

A second limitation is that the RVITM model provides an effective intensity-domain approximation rather than an exact description of coherent double-pass propagation. Residual nonlinearities arising from modal interference may therefore limit reconstruction fidelity, particularly for complex or highly structured targets. Model-based or physics-informed deep-learning approaches, in which the forward operator is explicitly incorporated into an unrolled reconstruction architecture [32], could provide a route to modelling these effects more accurately while retaining the physics-guided prior.

Beyond robustness, the restoration stage itself can be strengthened. Because it is decoupled from calibration, the backbones compared here could be replaced by stronger image priors without altering the reflected-RVITM front end. Untrained priors such as the deep image prior [33], learned generative priors including diffusion models used as plug-in solvers for imaging inverse problems [34], and correlation-based approaches shown to generalise through scattering media [35] are all natural candidates; these are particularly attractive given that hybrid cross-dataset SSIM remained near, rather than substantially above, the training-free physics reference, indicating headroom for priors that generalise more strongly than the supervised backbones used here.

Finally, translation towards minimally invasive applications will require further advances in spatial resolution, field of view, contrast, and acquisition speed. Multispectral or multiwavelength reflected operators could provide additional tissue-specific contrast [20], while distal-tip translation and image montaging could extend the effective field of view [20, 36]. Characterising frame rate, working distance, and performance in realistic three-dimensional and biological environments will also be essential. Taken together, these developments would move the reflected-RVITM framework towards minimally invasive applications such as microneedle guidance and *in situ* tissue inspection through an ultrathin single-fibre probe.

## 5 Conclusion

We have presented a single-shot, reflection-mode, single-MMF imaging framework that combines an intensity-only reflected RVITM with an image-restoration network to recover images from a single backscattered speckle frame, without interferometry or phase retrieval. On matched datasets, the reflected-RVITM-guided models generally outperformed both the analytic initial reconstruction baseline and the same backbones trained to invert the raw speckle directly, with only one non-significant difference in SSIM noted in the Results Section. Notably, hybrid models trained exclusively on Fashion-MNIST generalised to unseen CIFAR scenes, achieving mean PCC values of 0.61–0.65 compared with 0.36–0.50 for direct learning, and also transferred successfully to real reflective objects. These results demonstrate that combining a reflected-RVITM physics prior with an image-domain restoration network provides a practical and and generalisable approach to reflection-mode MMF imaging following intensity-only calibration. Future work will focus on bending-robust calibration and stronger restoration priors, with the longer-term goal of enabling minimally invasive, *in vivo* single-fibre endoscopy.

## Data Availability

All data produced in the present study are available upon reasonable request to the authors.

## Author contributions

Z.Y.: Conceptualization (equal), Methodology (leading), Software (leading), Investigation (leading), Writing – Original Draft (leading). Writing – Review & Editing (supporting). F.H.: Methodology (supporting), Writing – Review & Editing (supporting). T.Z.: Supervision (supporting), Writing – Review & Editing (supporting). W.X.: Conceptualization (equal), Supervision (leading), Funding Acquisition (leading), Writing – Review & Editing (leading).

## Data availability

Code supporting this study is available at https://github.com/Zye3/reflection-mmf-rvitm-dl. Associated data will be released publicly via the same repository upon publication.

## Supplementary data

Supplementary videos showing frame-by-frame reconstructions of the real objects are available online (Video 1: metal ventilation grille; Video 2: needle; Video 3: printed letter).

